# Microenvironment changes in basal ganglia region correlates with hypertension at 7T MRI

**DOI:** 10.1101/2023.10.05.23296633

**Authors:** Hongqin Liang, Shan Meng, Yue Li, Xiaoqi Yi, Haipeng Zhang, Jian Wang, Fajin Lv

## Abstract

**BACKGROUND:** Hypertension, a major risk factor for stroke, is associated with the changes of microenvironment in basal ganglia region, including microstructural characteristics and mapping relationship of blood flow and peripheral nerve tissue.

**MATERIALS AND METHODS:** The morphological characteristics of the lenticulostriate arteries (LSA) and peripheral nerve nuclei were compared by 7T time-of-Flight MRA between 32 hypertensive patients (mean age: 47.1±9.6 years) and 48 age-matched healthy subjects (mean age: 47.7±8.1 years) from September 2022 to February 2023. The ITK-SNAP software was used to measure the morphological characteristics of the LSA and the volume of the nuclei.

**RESULTS:** The average stems number and length of LSA was significantly higher in healthy people than those in hypertension patients (p<0.05). The volumes of the peripheral nerve nuclei in healthy subjects were significantly larger than those in hypertensive patients (p<0.05). There was a positive correlation between the volume of putamen and the length of LSA in the left hemisphere (r = 0.23, p < 0.05). The number of stems was found to be positively correlated with Globus pallidus volume (r = 0.27, p < 0.05) and internal capsule volume (r = 0.25, p < 0.05) in the right hemisphere. For older adults (>45 year old), the average stems number of LSA and the volume of nuclei were significantly different (P <0.05).

**CONCLUSION:** The microstructure of the basal ganglia has changed obviously in patients with hypertension compared to healthy individuals when observed by 7T MRA.

**NOVELTY AND RELEVANCE:** *What Is New?:* Previously studies on basal ganglia of hypertension using 7T MRI mainly focused on the imaging of LSA, but have not identified significant associations between the LSA and peripheral nerve tissue. However, the occurrence of hypertension adverse events such as intracerebral hemorrhage is closely related to the microenvironment formed by the lenticular artery and peripheral nerve tissue. Once the potential correlation is taken into account, the treatment and follow-up plan related to hypertension may be changed to effectively reduce the adverse.

*What Is Relevant?:* The rupture of LSA is a recognized cause of stroke in the basal ganglia of hypertensive patients, and the change of the microenvironment of peripheral nerve tissue is closely related to the rupture of LSA. The current findings confirm this notion and identify a potential association between the LSA and peripheral nerve nuclei in hypertensive patients, reinforcing the causal relationship between changes in follow-up strategies for lifelong hypertension and disease risk.

*Clinical/Pathophysiological Implications?:* Consider early and active assessment of basal ganglia blood flow and peripheral nerve tissue changes in all hypertensive patients to help reduce the risk of adverse events (such as cerebral hemorrhage, cerebral infarction, cognitive impairment, etc.) in individuals.

## INTRODUCTION

Hypertension is the most significant risk factor for stroke, also independent risk factors of cerebrovascular disease.^1^ The incidence of hypertension cerebral hemorrhage can be as high as 70% ∼ 80%.^2–4^ The basal ganglia is the most common site of hypertensive intracerebral hemorrhage, which accounts for the majority of subtypes of intracerebral hemorrhage. This type of hemorrhage often occurs when small blood vessels rupture, especially branches of the lenticulostriate artery (LSA).

From an anatomical and functional perspective, neural tissues in the basal ganglion are in contact with LSAs and their branches to maintain brain function.^5^ Therefore, LSAs and the neural tissues in the basal ganglion have a mutually interactive and affected relationship. This entity exhibits unique hemodynamic characteristics, which are key factors in the pathophysiology of LSA rupture Studies have shown that there is a significant pressure gradient between the LSA and peripheral nerve tissue, which increases the risk of LSA rupture.^6, 7^ Early visualization and quantitative evaluation of structural changes in LSAs and peripheral nerve nuclei in hypertensive patients effectively prevents basal ganglia cerebral hemorrhage.

Currently, conventional imaging methods cannot effectively evaluate the fine structure of LSAs and peripheral nerve nuclei due to low resolution. 7.0T magnetic resonance imaging (MRI) offers ultra-high signal-to-noise ratio and spatial resolution, enabling effective visualization of the microstructure of LSAs and peripheral nerve nuclei. Previous studies have mainly focused on small vascular diseases, diabetes and ischemic stroke.^8–11^ These findings suggest that 7T magnetic resonance angiography (MRA) can be used as a first-line diagnostic criterion for LSAs and peripheral nerve, especially in patients allergic to contrast agents and in follow-up patients. ^12, 13^ Previous studies mainly focused on the imaging technology of the lenticular artery. It is still unclear whether the lenticular artery and peripheral nerve tissue in hypertensive patients have microstructure changes at the same time, whether there is a correlation between the two and what the specific relationship is.

Therefore, we hypothesized that both LSAs and peripheral nerve nucleus microstructure changes were present in hypertensive patients, and there is a certain degree of correlation between the two. The primary aim of this study is to validate this hypothesis and report our findings on the structural differences between LSAs and peripheral nerve nuclei in hypertensive patients through quantitative analysis of 7T MRA.

## METHODS

### Study Design

This prospective study was a complete and balanced block design and approved by the institutional review board. Written informed consent was obtained from all participants. Thirty-two recently (within 2 weeks) diagnosed hypertensive patients and forty-eight participants of age and gender matched healthy volunteers as controls were enrolled consecutively. Hypertension was defined as a systolic/diastolic blood pressure of 140/90 mmHg according to 2018 ESC/ESH Guidelines for the management of arterial hypertension.^14^

They were included if they (a) had no abnormal brain lesions were found by obtaining anatomic images using a conventional 3.0T MRI (Siemens, Trio), (b) had taken no antihypertensive medications until this study. The exclusion criteria were as follows: (a) clinical contraindications to MRI (such as individuals with pacemakers, certain types of metallic implants, or severe claustrophobia). (b) Give up during the MRI, and (c) image quality is poor.

### Magnetic resonance imaging

All the subjects were examined on a 7T MRI (MAGNETOM Terra, Siemens Healthcare) equipped with a head gradient insert coil (80mT/m, 200T/m/s) and a 32-channel head coil (Nova Medical). TOF-MRA and T1-weighted magnetization-prepared rapid gradient echo (T1w-MPRAGE) sequences were collected for every subject. The T1w-MPRAGE was obtained for structural images, with the following parameters: FOV = 224 × 224 × 179 mm ^3^, resolution = 0.70 × 0.70 × 0.70 mm^3^, TR =3000ms, TE = 3.23ms, inversion time = 1050ms, FA = 8^◦^, BW= 320 Hz/Px, acceleration factor (generalized auto-calibrating partial parallel acquisition, GRAPPA) = 3, acquisition time (TA) = 5min54 sec. The imaging parameters of TOF-MRA were FOV= 180 × 135 × 52 mm^3^, resolution= 0.23×0.23×0.36mm^3^, TR= 15ms, TE= 3.57ms, FA= 20^◦^, BW=151Hz/Px, GRAPPA factor = 2, TA = 8min 20sec. The imaging slab was placed in an obliquely axial covering the upper bound of the basal ganglia and the lower bound of the Middle cerebral artery (MCA). TOF-MRA images were reformatted into maximum intensity projections (MIP) (projection thickness=25mm) coronal planes.

### Image analysis

Images were reviewed and analyzed using commercial soft-ware (ITK-SNAP). Coronal maximum intensity projection (MIP) with a slab thickness of 25mm was used to reconstruct the image for optimal displaying of LSAs. Tracing was manually performed by two radiologists with 5–7 years’ experience who recursively screened the MinIP/MIP images and delineated the center points along each visible arterial segment. The stem number, branch number, full length from the origin to the terminal, and local length (5-15mm from MCAs) of skeletonized vascular tree were analyzed.^15^ The generated LSAs skeletons were examined by another senior radiologist (with 10–15 years of experience in MRI image interpretation), ensuring the accuracy of the LSAs centerline. The stem is defined as LSAs originating directly from the first segment of the middle cerebral artery (MCA) (M1) or the anterior cerebral artery (A1). Branches were defined as daughter vessels arising from parent LSA stems without any single vessels as described previously. The maximum length was measured on the prominent LSA in the coronal MIP, which was the curved length from MCA to the visible end. The number of stems and branches and the maximal length were measured in each subject. The schematic demonstrates the method used for measuring the length and branch of the LSAs. The volumes of basal ganglia (BG) including putamen, caudate, internal capsule, thalamus, and Globus pallidum were measured on the T1w-MPRAGE by using ITK-SNAP. The volumes of basal ganglia (BG) morphological quantification workflow begin with manual vessel segmentation using ITK-SNAP on the raw TSE-VFA image. The volumes are reconstructed, and a mesh surface is created in preparation for shape analysis and quantitative measures. The specific quantification process of the lenticular artery and peripheral nerve nuclei was shown in figure 1.

**Fig.1.**
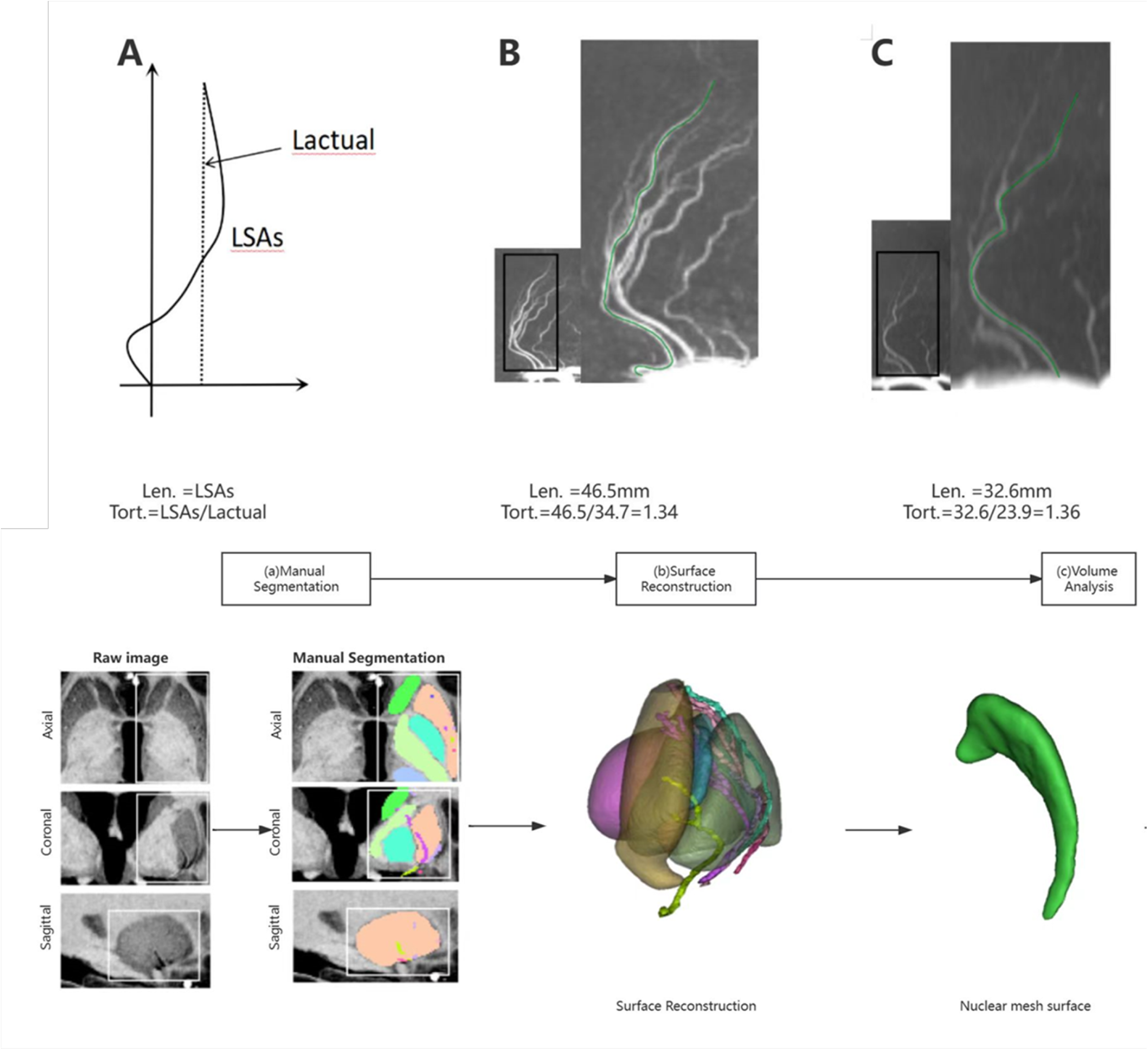
Methodology for measuring the length and tortuosity of the LSAs and the quantification workflow of basal ganglia nucleus. A Schematic illustration of the measurement of LSAs parameters. B, and C, Representative 7T MRA images of the LSAs, their tracings, and measured values for normal and hypertensive subjects, respectively. The basal ganglia nucleus quantification workflow begins with manual segmentation of the original image using ITK-SNAP (a). Reconstruct the surface and create a mesh surface to prepare for shape analysis (b). Quantitative measurements were made from the volume analysis diagram (c).

### Statistical analysis

Continuous variables were reported in the form of means±standard deviations (SDs), whereas a total number (percentages) were used for categorical variables. The independent t test was used to compare the morphologic characteristics of LSA and the volumes of basal ganglia nuclei on each hemisphere of the hypertensive and healthy groups. It was also performed to compare different age groups in subgroup analysis. The comparison of younger and older subjects in each group for LSA characteristics was illustrated by box plots. Statistical significance was defined as P<0.05. Partial correlation analysis was used to examine the relationship between morphological measurements of LSA and volumes of subcortical nuclei. All statistical analyses were performed by using commercial software (SPSS 26, IBM). Bonferroni correction was applied to the statistical results to reduce type-1 errors caused by multiple comparisons.

### RESULT

A total of 80 volunteers were included in analysis with acceptable image quality. The ages ranged from 34 to 69 years old. The demographics of participants in this study were showed in Table 1.

**TABLE 1.**
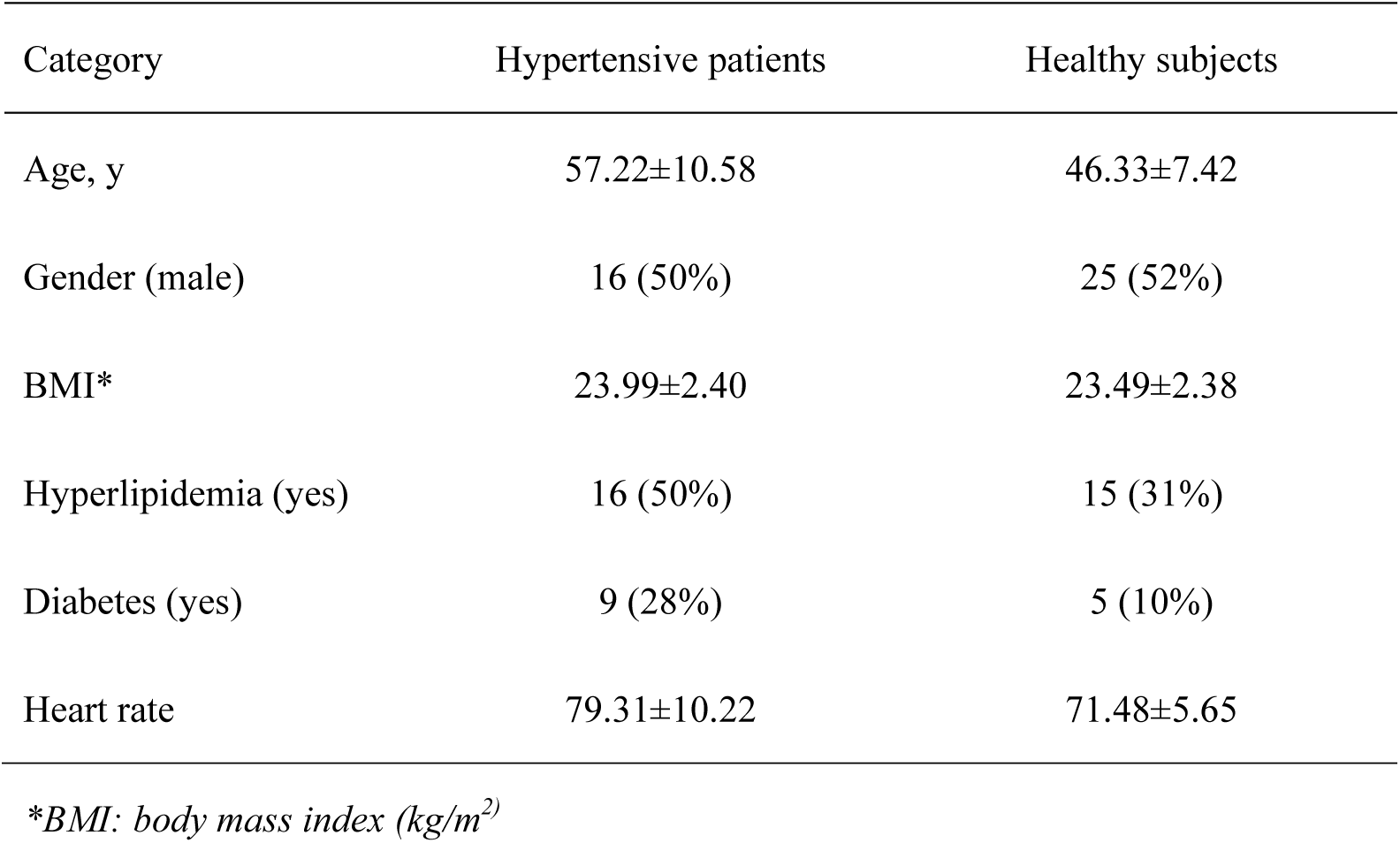
Demographic characteristics of hypertensive patients and healthy subjects.

| Category | Hypertensive patients | Healthy subjects |
| --- | --- | --- |
| Age, y | 57.22±10.58 | 46.33±7.42 |
| Gender (male) | 16 (50%) | 25 (52%) |
| BMI* | 23.99±2.40 | 23.49±2.38 |
| Hyperlipidemia (yes) | 16 (50%) | 15 (31%) |
| Diabetes (yes) | 9 (28%) | 5 (10%) |
| Heart rate | 79.31±10.22 | 71.48±5.65 |

\*BMI: body mass index ( $\text{kg/m}^2$ )

There were significant differences in stems and length by comparing the characteristics of LSA between hypertensive and healthy groups. The average number of LSA stems in the right hemisphere was significantly higher in healthy people than in hypertension patients (p = 0.013). In both hemispheres, the mean length of LSA in the healthy group was longer than in the hypertensive group (p = 0.011; p = 0.020). In both hemispheres, the volumes of the caudate nucleus, putamen, internal capsule, thalamus, and Globus pallidus in healthy subjects were significantly larger than those in hypertensive patients (Table 2).

**TABLE 2.** the characteristics of LSA and the volumes of basal ganglia nucleus between the hypertensive and healthy groups in both hemispheres.

|  | Left hemisphere |  |  | Right hemisphere |  |  |
| --- | --- | --- | --- | --- | --- | --- |
|  | Hypertensive patients | Healthy subjects | P value | Hypertensive patients | Healthy subjects | P value |
| LSA Stem (n) | 2.84±1.02 | 2.83±1.02 | 0.964 | 2.28±0.92 | 2.94±1.24 | 0.013 |
| LSA Branch (n) | 8.72±1.99 | 8.58±2.53 | 0.800 | 8.56±2.50 | 9.10±3.49 | 0.451 |
| LSA Length (mm) | 15.77±3.87 | 19.26±8.04 | 0.011 | 15.67±4.32 | 18.77±7.31 | 0.020 |
| Caudate nucleus (mm<sup>3</sup>) | 3155.11±346.07 | 3426.57±514.30 | 0.006 | 3151.58±354.26 | 3444.76±509.83 | 0.003 |
| Putamen (mm<sup>3</sup>) | 3952.20±442.02 | 4356.43±560.87 | 0.001 | 3932.89±426.17 | 4311.11±589.63 | 0.001 |
| Internal capsule (mm<sup>3</sup>) | 2061.42±393.90 | 2411.58±466.36 | 0.001 | 2049.87±393.57 | 2408.70±475.63 | 0.001 |
| Thalamus (mm<sup>3</sup>) | 4140.35±454.26 | 4559.01±636.78 | 0.001 | 4196.75±433.95 | 4560.55±572.94 | 0.003 |
| Globus pallidus (mm<sup>3</sup>) | 671.66±160.91 | 986.55±230.06 | 0.000 | 679.51±166.29 | 936.66±238.00 | 0.000 |

To investigate the influence of age on LSA characteristics and basal ganglia sizes, we divided hypertensive and healthy subjects into two groups: younger individuals (≤45 years old) and older individuals (>45 years old). The young hypertensive group only had significantly smaller putamen in the right hemisphere compared to young healthy group (P=0.009). However, the older group had significant differences in the average number of LSA stems in the right hemisphere (P=0.004). At the same time, there were significant statistical difference among the internal capsule, thalamus, and Globus pallidus in the left hemisphere (P<0.001, P=0.038, and P<0.001, respectively), as well as caudate nucleus, internal capsule, and Globus pallidus in the right hemisphere (P=0.048, P<0.001, and P<0.001, respectively). The distribution of whole-brain LSA stems, branches, and length among different age groups in hypertensive patients and healthy people. For the healthy group, subjects who were over 45 years old had a larger number of LSA stems than those who were under 45 years old (P<0.05). The average LSA length in the whole brain was longer in the healthy group than in the hypertension group (P<0.01). (Figure 2)

**Fig 2.**
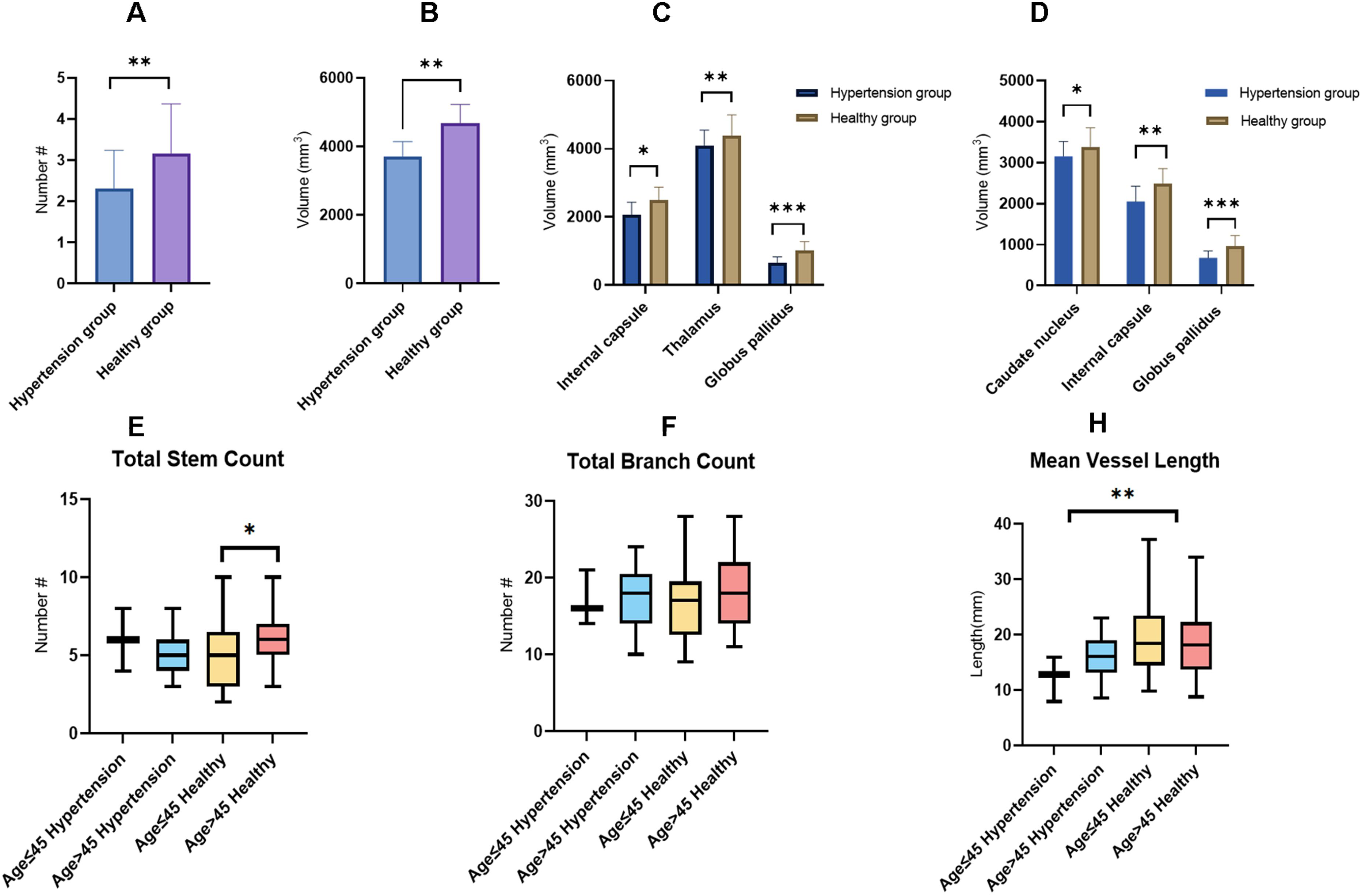
Comparison of basal ganglia volumes and LSA characteristics between the hypertensive and healthy groups. Average number of stems (A) and Average volumes of putamen (B) in the right hemisphere between hypertensive and healthy groups under 45 years old. Average volumes of internal capsule, thalamus and Globus pallidus in the left hemisphere between hypertensive and healthy groups (C) and Average volumes of caudate nucleus, capsule internal, and Globus pallidus (D) in the right hemisphere between hypertensive and healthy groups over 45 years of age.

After controlling for age, gender, the relationship between LSA morphology and subcortical nuclei was assessed. There was a positive correlation between the volume of putamen and the length of LSA in the left hemisphere (r = 0.23, p < 0.05). The number of stems was found to be positively correlated with globus pallidus volume (r = 0.27, p < 0.05) and internal capsule volume (r = 0.25, p < 0.05) in the right hemisphere. (Figure 3)

**Fig 3.**
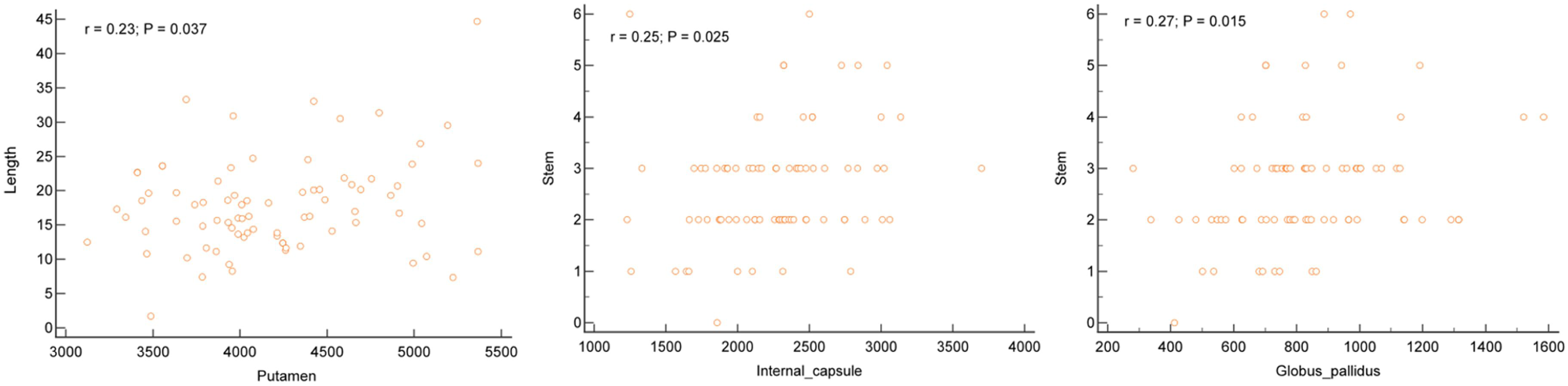
The correlation between LSA and nerve nuclei. A, Correlation between LSA length and putamen volume in the left hemisphere; B, Correlation between LSA stems and internal capsule volume in the right hemisphere; C, Correlation between LSA stems and globus pallidus volume in the right hemisphere.

## DISCUSSION

In this study, we transitioned from a simple LSA study to investigating the microenvironment formed by LSA and the peripheral neural nucleus. Using 7T TOF and MPRAGE technology, we have obtained more detailed information for the nuclei and the morphological characteristics of LSA in hypertensive patients and age-matched healthy people, which has been challenging at conventional imaging techniques. We found the microenvironment of blood flow and nerve tissue formation in basal ganglia has undergone microstructural changes in hypertensive patients, that is, the number, branches of LSA and nuclear mass volume of the basal ganglia were reduced compared with the healthy group, and this change was especially pronounced in the elderly.

Our findings are consistent with some previous studies in which the number and length of LSAs visualized for hypertensive patients were significantly less than those in age-matched healthy population from 7T TOF-MRA. There are several possible explanations for this. First, LSAs are a group of tiny arteries consisting of a single branch originating in the horizontal segment of the middle cerebral artery, with an outer diameter of only 0.08-1.4mm.^16, 17^ As everyone knows, hypertension is the primary cause of microvascular structural damage. It can accelerate arteriosclerosis, weaken the elasticity of blood vessels, or their ability to dilate, and leads to narrowing of the lumen, which cannot reach a resolution of 7T MRI.^18^ In addition, the increased peripheral vascular resistance caused by hypertension will lead to decreased microvascular blood flow and consequent decrease in blood flow signal strength, which may make it difficult for microvessels to be shown in angiography.^18,19^ Meanwhile, we also found that the visualization of lenticular artery in hypertensive elderly patients was significantly lower than that in age-matched healthy people. Previous research reported the decreased flow velocity of LSA was found in the elderly by a phase-contrast MRI study, which appears to reflect the degeneration changes of the aging cerebrovascular system.^20, 21^ and hypertension accelerates the process of degeneration and arteriosclerosis, including the weakening of vascular elasticity, contractility, and local hemodynamic changes.

Studies have found that patients with hypertension often show volume changes in the basal ganglia region, including atrophy or volume reduction of multiple nerve nuclei.^22,23^ Previous literature also reports that the atrophies of the BG and other subcortical nuclei have been happened in various neurodegenerative diseases.^24^ In our study, the volume of the caudate nucleus, putamen, internal capsule, thalamus and pallidum in the basal ganglia region was significantly smaller in hypertensive subjects than those in healthy patient. This is consistent with previous studies and may be related to the damage or death of basal ganglia nerve cells caused by insufficient blood supply, oxidative stress and inflammation caused by hypertension.^25, 26^ In addition, previous studies have found that both the volume of BG are found to decrease with age in healthy volunteers.^27, 28^ In this study, we found that this phenomenon is not only in healthy people, but also in patients with high blood pressure. These findings are reasonable, and this may be because with the increase of age and the continuous rise of blood pressure, hypoxia caused by reduced blood flow and abnormal neurotransmitter transmission caused by continuous resistance led to obvious atrophy of the nerve nuclei in the basal ganglia. ^29^

In addition, we found that there was a certain correlation between the morphology of the lenticularis artery and some peripheral nerve nuclei. There was a positive correlation between the volume of putamen and the length of LSA in the left hemisphere, while the number of stems was found to be positively correlated with globus pallidus volume and internal capsule volume in the right hemisphere. Previous literature shows that the volume of putamen was associated with the number of LSAs, while the volume of caudate was closely related to the length of LSAs in the healthy.^27^ In our study, we found that there were differences in the bilateral basal ganglia nuclei of hypertension, which were correlated with the LSA, which may explain why cerebral hemorrhage in the basal ganglia is mostly unilateral and has different symptoms. All of it again shows the unity of the blood flow in the basal ganglia and the nerve nucleus mass to jointly maintain the function of the brain region, which facilitate the assessment of the risk of cerebral hemorrhage in hypertensive patients.

There are several limitations in our study. Firstly, the complete basal ganglia microenvironment includes nerve nuclei, LSA and white matter fiber bundles. In this study, we did not focus on the network construction of white matter fiber tracts in hypertensive patients, mainly because it takes a lot of time to collect many image sequences at the same time, and the patient was unable to persist during the examination. We plan to conduct imaging and research on Diffusion Tensor Imaging (DTI) specifically in hypertensive patients in subsequent studies. Secondly, our study included a relatively small sample, and the next large-sample multi-center research will be one of our efforts in the future. Finally, advanced automatic segmentation and reconstruction techniques need to be further applied to better objectively measure fine structures.

In conclusion, we extended previous studies from simple LSA imaging to the quantitative assessment of the microenvironment of the LSA and peripheral nerve nuclei, making the changes between basal ganglia blood flow and neural nucleus microstructure in hypertensive patients as an organic whole, and providing additional evidence for unilateral morbidity and different prognosis in hypertensive patients. It provides important clues for evaluating the adverse effects of small vessel diseases and stroke in special population such as hypertension.

## PERSPECTIVES

The current data analysis shows that the microstructure of the lenticular artery and peripheral nerve tissue has been changed in hypertensive patients. The morphology (including total length, number of stems and branches) and the nucleus mass in basal ganglia were significantly correlated with age and blood pressure. There was a certain correlation between the LSA and different nucleus mass. These findings provide a compelling hypothesis that the LSA alone does not fully predict the occurrence of hypertensive adverse events. Therefore, it may be an important population health strategy to consider active management of the basal ganglia microenvironment in early stage in all adults with hypertension, not just individuals after adverse hypertensive events.

## Data Availability

The original contributions presented in the study are included in the article, further inquiries can be directed to the corresponding authors.

## Nonstandard Abbreviations and Acronyms

LSA: lenticulostriate artery
MRI: magnetic resonance imaging
MRA: magnetic resonance angiography
MIP: maximum intensity projections
MCA: middle cerebral artery
BG: basal ganglia
DTI: diffusion tensor imaging

## Acknowledgements

The authors acknowledge the volunteers who participated in the study.

## Author Contributions

Hongqin Liang, Shan Meng, Fajin Lv, and Jian Wang conceived the study and developed its design; Haipeng Zhang, Shan Meng and xiaoqi Li collected and processed data; Yue Li performed statistical analyses; Hongqin Liang wrote the first draft of the article; and all authors contributed to data interpretation and revision of the article.

## Ethical statement

This study was performed in line with the principles of the Declaration of Helsinki. Approval was granted by the Ethics Committee of Southwest Hospital, Third Military Medical University [Army Medical University (No. (A)KY2023071)

